# Integration of dilated cardiomyopathy genomics with transcriptomics from the human heart implicates regulatory molecular mechanisms

**DOI:** 10.64898/2026.01.12.26343934

**Authors:** Connor S. Murray, Chaojie Yang, Suet Nee Chen, Sharon Graw, Anis Karimpour-Fard, Joseph Cleveland, Shanshan Gao, Hae Kyung Im, Heather E. Wheeler, Amrut V. Ambardekar, Jordan R. H. Hoffman, Stacey Gabriel, Namrata Gupta, Kristin Ardlie, Jerome I. Rotter, Kent D. Taylor, Stephen S. Rich, Luisa Mestroni, Ani Manichaikul, Matthew R. G. Taylor

## Abstract

Heart failure (HF) is a leading global cause of morbidity and mortality, yet the regulatory molecular mechanisms that link genetic variation to cardiac dysfunction remain elusive. To bridge this gap, we created the Trans-Omics for Precision Medicine in Congestive Heart Failure (TOPCHeF) resource, a multi-omics dataset comprising >700 human left-ventricular tissue samples, including dilated cardiomyopathy (DCM), ischemic cardiomyopathy (ICM), and non-failing controls, with paired whole-genome and RNA sequencing. By mapping expression-(eQTL) and splicing-(sQTL) quantitative trait loci directly in diseased human hearts, we identified over 10,000 transcripts with significant eQTL and 8,600 isoforms with significant sQTL, across both coding and non-coding genes, many of which overlap loci previously associated with HF and emerging novel gene associations. Single-locus colocalization with a largescale DCM genome-wide association study revealed 21 expression and 17 splicing-QTL that share causal variants with disease risk. These include known Mendelian cardiomyopathy risk genes such as *FLNC* and *ACTN2*, and novel regulatory candidates like *CAMK2D*, *LMF1*, *MYOZ1*, *SKI*, *SYNPO2L*, and *TKT*. Several loci also showed coordinated effects on both gene expression and RNA splicing, implicating calcium signaling, cytoskeletal organization, and metabolic pathways in HF pathogenesis. Together, these results help define the regulatory landscape of the failing human heart and establish TOPCHeF as a foundational resource for connecting genetic variation to transcriptional and splicing molecular mechanisms in HF research.

## Introduction

Heart failure (HF) is a major public health challenge and a leading cause of morbidity and mortality worldwide. Over 6 million Americans have HF (Bozkurt et al., 2023) and is the leading cause of death for over 860,000 yearly (Tsao et al., 2023). In 2021 alone, HF was estimated to cost over $284 billion worldwide through direct and indirect costs, leading to a large economic strain on the global healthcare system (Darvish et al., 2025). HF is a diverse clinical disorder that encompasses multiple disease states (Groenewegen et al., 2020). Among these, dilated cardiomyopathy (DCM) is characterized by left ventricular (LV) enlargement and reduced contractile function that is not explained by coronary artery disease or other loading conditions (e.g., hypertension and valvular heart disease; Schultheiss et al., 2019). While ischemic cardiomyopathy (ICM) is defined by LV dysfunction in the presence of significant coronary artery disease (Pastena et al., 2023). Alongside biospecimens obtained from individuals with DCM and ICM, samples from non-failing hearts provide a critical source for comparison in investigations of disease-specific molecular mechanisms of HF.

Advances in functional genomics and high-throughput sequencing offer new opportunities to interrogate disease mechanisms across tissues (Wray, 2007). Expression quantitative trait loci (eQTL) and splicing quantitative trait loci (sQTL) link genetic variation (single nucleotide polymorphisms; SNPs) to transcriptional (mRNA abundance; Cheung et al., 2003) and post-transcriptional regulation (alternative intron excision and splice junction usage; Li et al., 2018; Qi et al., 2022), respectively, and have proven invaluable for identifying disease causal variants across myriad examples. For instance, coronary artery disease (Liu et al., 2018; Yao et al., 2018), atrial fibrillation (Liu et al., 2021), and myocardial infarction (Zhou et al., 2025) have all been investigated with both diseased and non-diseased tissues in exploratory approaches. Colocalization analyses, which combines QTL with genome-wide association study (GWAS) signals enable prioritization of putative causal genes and variants (Giambartolomei et al., 2014; Guo et al., 2015).

The non-diseased heart has been investigated for its QTL landscape (Gupta & Musunuru, 2013; Koopmann et al., 2014), including the atrial appendage and LV from the Genotype-Tissue Expression (GTEx) project that uncovered that nearly every gene has regulatory associations across both eQTL and sQTL (The GTEx Consortium et al., 2020). However, it remains uncertain whether QTL derived from diseased tissues (DCM and ICM LVs) can provide greater interpretive power for GWAS loci than those derived from large population-based reference cohorts. The Myocardial Applied Genomics Network (MAGnet) consortium performed eQTL discovery using array-based measurement of gene expression across 313 humans hearts (177 failing, 136 non-failing donors), finding an enrichment of histone modifications across both non-failing and failing cohorts (Cordero et al., 2019). While some work has been addressed on diseased heart tissues, there are limited resources available for HF outside of GTEx and MAGnet to serve for QTL replication and further tissue-specific investigation.

High-impact genetic variants have been implicated in cardiomyopathies, including Mendelian risk alleles in genes such as *Filamin C* (*FLNC*), *BAG family molecular chaperone regulator 3* (*BAG3*)*, alpha-actinin-2* (*ACTN2*)*, Prelamin-A/C* (*LMNA*), and *Titin* (*TTN*) (Rasooly et al., 2023; Shah et al., 2020; Tadros et al., 2025). Yet, cardiomyopathy is a complex disease state with polygenic origin outside of large effect alleles (Jacoby & McKenna, 2012), DCM alone routinely has 40-50 genes used by commercial testing laboratories for disease evaluation (McNally & Mestroni, 2017). These large-effect mutations illustrate the critical role of both common and rare variants in shaping HF susceptibility and underscore the need for systematic approaches to connect genetic risk to molecular mechanisms and refining etiology.

To address these concerns, we introduce the Trans-Omics for Precision Medicine in Congestive Heart Failure (TOPCHeF) project, a resource comprising >700 adult donor left-ventricle heart samples from which both RNA and DNA were extracted and sequenced. TOPCHeF is uniquely positioned to advance exploratory and prioritization genetics research in HF. By including DCM, ICM, and non-failing hearts, we provide an unprecedented opportunity to evaluate eQTL and sQTL across failing and non-failing states, thereby informing future validation efforts for gene-editing experiments. Using this experimental framework, we identified over 10,000 significant eQTL and ∼8,600 isoforms with sQTL, of which colocalization with a large DCM GWAS (Jurgens et al., 2024) revealed 21 and 17 colocalized genes, respectively (Figure 1A-D). Among these colocalized loci, we highlight well-documented Mendelian DCM genes such as *FLNC* and *ACTN2* as well as emerging non-Mendelian candidates including *CAMK2D, LMF1, MYOZ1*, *SKI, SYNPO2L,* and *TKT*. Collectively, our work expands the regulatory landscape of genetic variation underlying HF biology and provides a foundation for exploratory precision medicine within the diseased heart’s regulatory variation (Lau & Wu, 2018).

**Figure 1:**
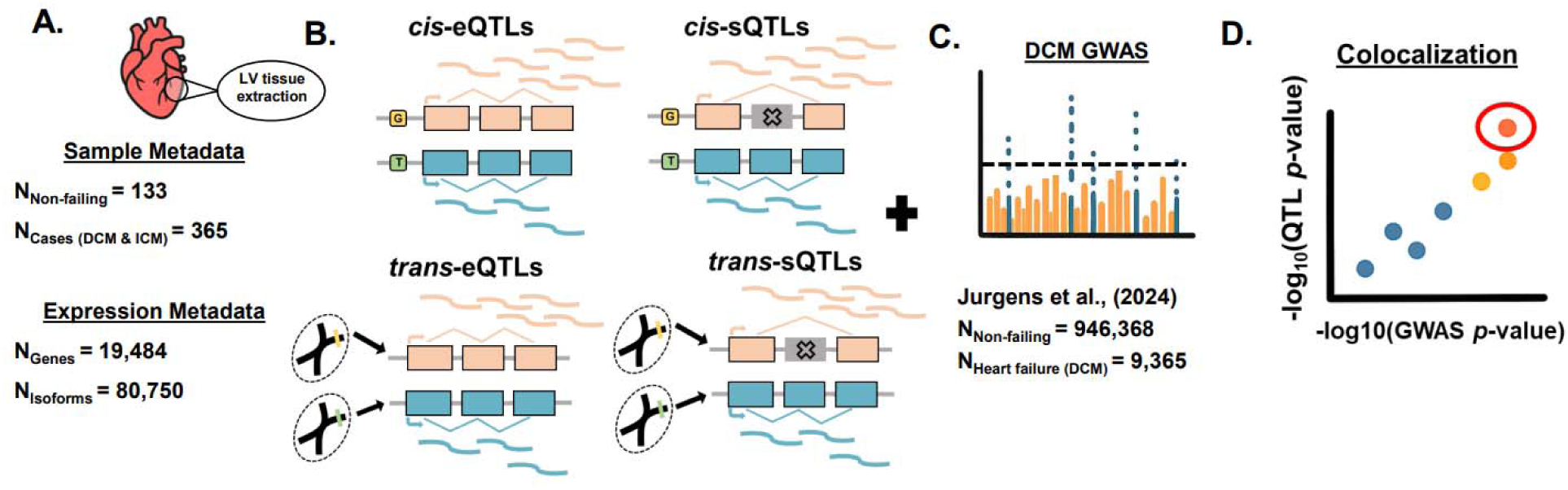
Introduction to the TOPCHeF dataset, quantitative trait loci (QTL) analyses, and variant prioritization. **A.** Metadata summary for left ventricular samples collected from failing and non-failing hearts, along with the number of filtered genes and isoforms included in eQTL and sQTL mapping, respectively. **B–D.** Schematic cartoon of the TOPCHeF analytical framework. **B.** Testing for eQTL and sQTL in both cis– and trans-contexts. **C.** Dilated cardiomyopathy (DCM) GWAS used for variant prioritization analyses with number of samples. **D.** Colocalization of cis-QTL and DCM GWAS signals for further prioritization.

## Materials and Methods

### Study Population and Enrollment

The TOPCHeF dataset is comprised of >700 sequenced individuals, of which 516 has paired-RNA and –DNA whole-genomes from adult hearts submitted to the TOPMed consortium Freeze 10b (Taliun et al., 2021). General demographics metadata are in Table 1 along with disease diagnoses. Adult patients undergoing cardiac transplantation at the University of Colorado are invited to provide informed consent for research participation in the cardiac tissue biobank, which has been an ongoing protocol for the past three decades. Clinical phenotype information on the etiology of end-stage HF is extracted from the medical records along with basic demographic data including age, sex, race, and ethnicity. When possible, clinical diagnoses and phenotype information are extracted from clinical notes of HF from cardiologists. DCM samples are from patients with advanced HF without other known causes of HF (e.g. ischemic, viral, alcohol, or longstanding, untreated risk factors of diabetes, hypertension, hypercholesterolemia). ICM samples are classified based on history and/or documentation of ischemic events or confirmed coronary artery disease. Explanted heart tissue from diseased hearts is obtained at the time of transplant in the operating theater. Non-failing cardiac samples are obtained from organ donors whose hearts were unsuitable for transplantation due to factors such as size or ABO blood group mismatch, with consent provided by family members via the local organ procurement agency. In this study, LV tissue was studied and only DCM, ICM, and non-failing samples were considered.

**Table 1:**
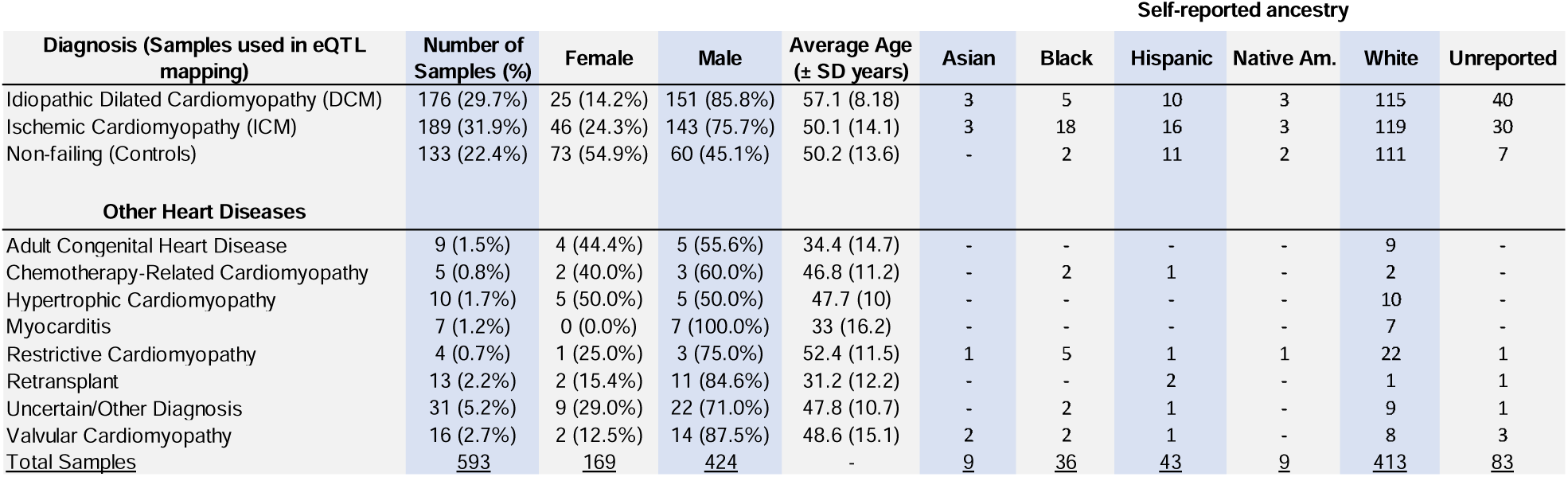
Demographic metadata of TOPCHeF genomes. All samples are from adult tissues. Self-reported ancestries were gathered during patient visits. The self-reported ancestries do not serve any functional use as covariates within this work and largely serves as a qualitative indication of the genetic diversity captured by *TOPCHeF*. Dashes indicate empty variables and the “Unreported” category represents any samples either without self-reported ethnicity and race information or were optionally withheld.

### Sample Procurement and Preparation

Upon explantation, approximately one to five gram aliquots of LV free wall tissue, visibly remote from scarring or infarction, are obtained and immediately frozen in liquid nitrogen, transported to the research lab, and stored, long-term, at −80 °C. Clinical samples from other cardiac regions have been increasingly collected in recent years and were not included in the current study. Clinical characteristics of tissue are annotated in a secure REDCap database and can be connected to whole genome and proteome data on the samples. Genomic DNA was extracted directly from LV tissue using Qiagen DNeasy Blood & Tissue Kits (Cat no. / ID. 69504). Whole genome sequencing of samples that met quality control metrics was completed by the Broad Institute. Mapping, alignment, and variant calling was done using the Illumina DRAGEN pipeline, https://support.illumina.com/content/dam/illumina-support/documents/documentation/software_documentation/dragen-bio-it/dragen-bio-it-platform-v3.2.8-user-guide-1000000085871-00.pdf. RNA was extracted from LV tissue (∼30 mg) that was homogenized. RNA extraction was done using Qiagen RNeasy Mini Kits (Cat no. / ID. 74104). RNASeq of poly-A enriched extracted high-quality RNA was performed by the Broad Institute of MIT and sequenced using the Illumina HiSeq 4000 (Illumina, San Diego, CA) platform, for a target depth of ≥ 40 M 2 × 101 bp paired-end reads. Alignment was performed using the TOPMed RNA-seq pipeline. Reads were aligned to GRCh38 with STAR (Dobin & Gingeras, 2015) and collapsed to the gene-level using RNASEQC v2 (Graubert et al., 2021) and the GENCODE 34 reference. Further pipeline details are available at: https://github.com/broadinstitute/gtex-pipeline/blob/master/TOPMed_RNAseq_pipeline.md.

### A Nextflow Computational Pipeline for QTL Mapping and Colocalization Analyses

We have built a *nextflow v23.04.1* pipeline (Di Tommaso et al., 2017), to study the cis and trans-e/sQTL given paired RNA– and DNA-WGS data from both diseased and control patient cohorts. *Nextflow* is a workflow manager that streamlines the “flow” of data between processes and successive jobs and is transferable to both cloud-based and high-performance computing environments. We describe below each component of the pipeline and present a graphical representation of processes in Supplemental Figure 1 and the software can be accessed here: https://github.com/connor122721/nf-eqtls.

### Genetic Variants Quality Control and Filtration

We downloaded the Freeze 10b TOPMed VCF (839 M SNPs) and subset to participants of the TOPCHeF study by using *bcftools v1.17* (Danecek et al., 2021). We first removed any sites that had a minor allele count < 1, insertions or deletions (indels) ≥ 50 bps, and sites that did not pass the SNP filters imposed by TOPMed (Taliun et al., 2021). In total, we had genotype data for 44,107,358 SNPs that passed all initial filtration steps. For analysis that excludes any sites with a minor allele frequency < 0.01, we retain 43.93M SNPs.

### Kinship Analysis of Related Individuals

We used *KING v2.3.2 ––related* (Manichaikul et al., 2010) on the filtered VCF to test if any TOPCHeF samples are related. We identified 19 full-sibling pairs and 2 first degree cousins and excluded one random sample within each pair, so their relatedness does not confound further analyses. Subjects were enrolled consecutively and without any selection based on disease phenotype or family history. Based on patient access in the Rocky Mountain region, some instances of biological relatives like sibling pairs, parent-offspring, and other family relationships occurred in the enrollment process.

### Genome-wide Variant Principal Component Analysis (PCA)

We tested how samples were clustered by principal components (PC) of ancestry using *snpgdsPCA()* in *SNPRelate v1.36.1* (Zheng et al., 2012). We first removed SNPs with a minor allele frequency (MAF) < 0.01, adjacent SNPs in high linkage disequilibrium (*R^2^* > 0.1, 100 bp windows, 10 bp step size) using *plink v1.9* (Chang et al., 2015), and thinned every 250^th^ SNP using *VCFtools v0.1.16* (Danecek et al., 2011). We retained 161,675 SNPs for use in PCA after filtering and converted the VCF to a genomic data structure using *SeqArray v1.42.4* in *R* (Zheng et al., 2017). The PCA shows a structure based on self-reported ethnicity, specifically the first PC accounts for an African (i.e., Black) and non-African (i.e., White, Hispanic, Asian, Native American) group, while PC2 tends to separate Asian and Hispanic background samples (Supplemental Figure 2). Yet, it should be noted that self-reported ethnicity does not always match genetic ancestry.

### RNA Normalization, Filtration, and PCA

We analyzed gene-level read counts from the TOPCHeF dataset, which included 58,103 genes (i.e., protein-coding and non-coding genes) before filtering. Gene read counts were imported into *R*, and we applied median ratio size-factor normalization (Anders & Huber, 2010) using *DESeq2 v1.42.1* (Love et al., 2014). Genes with normalized coverage below 10 across at least 10 samples were excluded from the dataset. Additionally, we applied group-specific filtering to remove genes with low normalized coverage (< 10) in at least 10 samples within the HF group (i.e., DCM & ICM samples), resulting in the removal of 151 genes. To control for potential biases, we identified and removed sex-biased genes by performing an analysis of variance test (ANOVA) on normalized gene counts by sex (i.e., Normalized Gene Count ∼ Sex), removing 241 genes that showed statistically significant sex-differences in expression after Bonferroni *p*-value correction. We also excluded ERCC-spiked genes and retired gene models. For further preprocessing, zero counts of genes in the dataset were replaced with half the minimum observed non-zero count for each gene, followed by a log_10_ transformation.

Principal component analysis was conducted on the normalized and filtered gene count matrix using *PCA()* in *FactoMineR v2.11* (Lê et al., 2008). Outliers in PCA space were identified using two approaches. First, we flagged samples as outliers if their mean absolute deviation (MAD) across the first 10 PCs, normalized by the dataset’s MAD, was ≥ 7. Second, we calculated the Mahalanobis distance of the first 5 PCs and classified samples as outliers if their Bonferroni-corrected *p*-value from a chi-square test was < 0.05. Based on these criteria, we identified 13 outliers, with all but one sample overlapping between the two RNA outlier detection methods. Biological sex was inferred using the expression of *XIST* (chrX; *ENSG00000229807*) and *RPS4Y1* (chrY; *ENSG00000129824*) for females and males respectively. This analysis revealed no ambiguous or cross-sex contaminated samples (Supplemental Figure 3).

### Cell-type Deconvolution for Bulk RNA-seq

Cell-type composition can both influence and reflect disease states, and single-cell RNA sequencing provides a powerful way to test for such differences in case-control experiments of the heart (Koenig et al., 2022). Although all tissue in this study was collected from the LV, we sought to determine whether large shifts in cell states distinguish affected from control samples. To do this, we applied *MuSiC v2* (Fan et al., 2022) to estimate cell-type proportions from TOPCHeF bulk RNA-seq data, using the Heart Cell Atlas v2, a reference atlas of human heart single-cell RNA-seq (Kanemaru et al., 2023; Litviňuková et al., 2020). For the reference dataset, we retained only cells that were captured with scRNA-seq in the left-ventricular tissue, with at least 500 detected genes and more than 800 total reads, and restricted analysis to the following labeled cell classes: cardiomyocytes, endothelial cells, fibroblasts, neural cells, and mural cells. We chose these 5 cell-types due to their high relative frequency in the TOPCHeF dataset during initial deconvolution testing.

### Gene and variant mappability calculation

A concern for cis and trans-QTL mapping was that stretches of similar sequence across distinct regions of the genome will result in alignment errors from short read sequencing experiments. Alignment errors can result in the inflation of false positive signals and increase the burden for multiple testing correction, especially for trans-QTL which typically have lower effect sizes (Saha & Battle, 2019). We used the nextflow implementation of *crossmapp* (https://github.com/porchard/crossmap-nextflow) and the GENCODE v34 GTF, with exon kmer length set to 100 bps, UTR kmer length set to 36 bps, and allowing two mismatches to calculate a gene-level bed file for mappability scores (Orchard et al., 2025). We used the mappability scores for filtering during QTL mapping below.

### Expression Quantitative Trait Locus (eQTL) Mapping

To understand how common mutations (MAF > 0.01) are implicated in expression level (i.e., transcript abundance) of genes between cases and non-failing hearts, we ran *TensorQTL v1.0.10* (Taylor-Weiner et al., 2019) using the filtered SNP dataset with a flanking window of ± 1 Mbps and 10,000 permutations enabled to estimate permutation *p*-values. As an initial covariate model, we included sex, age, disease group (e.g., affected versus non-failing), and cardiomyopathy status (i.e., DCM & ICM), as well as RNA PCs, and the first 5 PCs of ancestry (i.e., DNA PCs). Where sex, disease group, and cardiomyopathy diagnoses are encoded as binary values (0,1). Using this general covariate model, we ran the *cis* mode to identify all significant eQTL for each gene based on a permutation *p*-value < 0.05 while varying the amount of RNA PCs as covariates from 1-100 (ex. RNA PC_1_ … RNA PCs_1-100_). We found that the covariate model that uses RNA PCs_1-70_ resulted in the most significant eGenes and is used for every follow up analysis of eQTL (Supplemental Figure 4A). Additionally, it appears that the saturation points of eGenes (e.g., statistical plateau) was reached somewhere between the inclusions of RNA PCs_1-50_ and RNA PCs_1-75_ (Supplemental Figure 4A&B). The *cis-nominal* mode of *TensorQTL* was ran to get pairwise-SNP summary statistics for use in colocalization analyses of significant candidate eGenes using the RNA PCs_1-70_ covariate model to ensure computational efficiency (see colocalization methodology below). The final covariate model we used to map *cis*-eQTL is as follows:

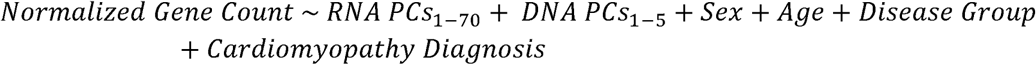

Variants were considered fine-mapped cis-variants if they appeared in the 95% credible sets output by TensorQTL SuSiE for the relevant cis-eQTL regions. We also investigated trans-eQTL, or distal loci influencing a gene’s expression, to understand long-range interactions between variants and gene expression profiles. We used the *trans* mode of *TensorQTL* using the covariate model listed above to estimate trans-eQTL (MAF > 0.01) and only retained significant regions based on a conservative genome-wide cut-off (*p* < 5 × 10^−8^). We summarized the count of significant eGenes and eVariants to see if there are any regions showing support for being hotspots and report on their trans-eQTL profiles.

### Splicing quantitative trait loci (sQTL) mapping

We assessed splicing-QTL (sQTL) by first running *regtools junctions extract v0.5.2* (Cotto et al., 2023) to get the counts of every exon-exon junction with a minimum intron length of 50 bps (*-m 50*), and an anchor length of 8 (*-a 8*) across each bam file. We then clustered the introns found among the junction files using the *leafcutter v0.2.9* (Li et al., 2018) toolset. We used the *cluster_prepare_fastqtl.py* script (https://github.com/broadinstitute/gtex-pipeline/blob/master/qtl/leafcutter/src/cluster_prepare_fastqtl.py), while considering any intron less than 500 kbps (*-l 500000*), any cluster with at least 10 reads (*--min_clu_reads 10*), and kept a cluster that had at least 0.001 fraction of reads supporting a junction (*--min_clu_ratio 0.001)*. We removed clusters with no counts in most of the samples, low complexity clusters, and normalized the matrix, and then each splicing phenotype was inverse normalized. This normalized matrix was used as input to the cis-sQTL scans, and splicing PCs were used as scan covariates. We also added gene information from the collapsed GENCODE GTF *v34*, and for each gene, variants within 1 Mbps of the gene’s TSS were tested. Gene TSS locations were determined using pyqtl’s gtf_to_tss_bed function. Trans-sQTL were tested with the same covariates as the cis-sQTL and restricted to MAF > 0.05 and only retained significant sites after a conservative Bonferroni correction based on the number of unique isoforms tested (α = 5 × 10^−8^ / 80,786).

### Colocalization of cis-QTL with DCM GWAS Summary Statistics

To prioritize disease-relevant cis-eQTL and cis-sQTL, we used colocalization to find overlap with a previously published HF GWAS. We implemented colocalization to test for a shared causal variant for GWAS and QTL in a ±1 mbp window of the genome under the assumption of one causal variant (Giambartolomei et al., 2014). We identified the Jugens et al., (2024) HF GWAS with large sample sizes (N_Control_ = 946,368; N_Cases_ = 9,365) to use for colocalization. Jurgens et al., (2024) identified 70 genome-wide significant loci associated with DCM across a multi-ancestry cohort. Briefly, the clinical state of DCM was identified in Jurgens et al., (2024) using the following metrics: 1) the left ventricular ejection fraction (LVEF) being less than 50%, 2) the left ventricular end-diastolic volume index (LVEDVi ≥ 75 mL/m^2^ for males, LVEDVi ≥ 62 mL/m^2^ for females), 3) the left ventricular end-diastolic diameter (LVEDD > 58 mm for males, and > 52 mm for females).

Using the GWAS summary statistics, we used the *coloc.abf()* function in the *coloc v5.2.3* package in *R* under default settings (Giambartolomei et al., 2014). We ensured the alternative and reference alleles were concordant between QTL and GWAS results and switched effect signs and alleles if there were any discordant sites. We standardized “effect alleles” as the alternative allele based on the human genome for each site. We identified a colocalized loci if the gene tested had a PPH.4 ≥ 0.8 for each gene window. For each colocalized region, we also calculated the pairwise LD (squared correlation, R²) among SNPs within the TOPCHeF cohort using *plink/1.9ob7.2*.

### Protein-protein interactions (PPI)

We used the *STRING v12.0* web browser tool (Szklarczyk et al., 2023) that combines several largescale protein binding databases and machine learning (e.g., random forest and neural nets) to predict protein binding partners when given an amino acid sequence. We used the default settings for all analyses.

### Statistics and Visualization

Most analyses were performed using *R v4.3.1* (R Core Team, 2013). We used the following packages for analysis and visualization: *tidyverse v1.3.1* (Wickham et al., 2019), *ggplot2 v3.3.5* (Villanueva & Chen, 2019), *patchwork v1.0.1* (Thomas Lin Pedersen, 2022), *data.table v1.12.8* (Dowle & Srinivasan, 2023), *foreach v1.4.7*, *doMC v1.3.5* (Daniel et al., 2022), *SeqArray v1.26.2* (Zheng et al., 2017).

## Results

### Heart failure whole-genome sequencing dataset

We present a novel dataset, TOPCHeF, to expand our understanding of the genetic and transcriptional basis of HF while also representing a valuable resource for other researchers studying HF (Figure 1A-D). TOPCHeF contains >700 samples that have paired RNA– and DNA whole-genome sequencing (WGS) available within the freeze 10b of TOPMed accessed on September 2024 (Figure 1A). We focus analyses on the DCM, ICM, and non-failing adult groups due to their large relative sample sizes representing 84% of total samples (Table 1). In DCM and ICM, the sex and age distributions are similar with a greater representation of male samples (range: 75-85%). The non-failing control group has more equal representation of male and female samples and is similarly aged with both DCM and ICM groups. Below, we delve into the genetic composition and ancestry components of the TOPCHeF dataset for DCM, ICM, and non-failing samples.

### Transcriptional and deconvoluted cell-type variation within the TOPCHeF dataset

Pairwise correlation of covariates with RNA PCs reveals the major sources of variation in the TOPCHeF transcriptomes were not closely related to the selected covariates including sex, age, and disease state (Supplemental Figure 5). However, RNA PC1 correlates generally (Pearson’s R^2^ > 0.2) with “container ID” (i.e., sample preparation container) and “mapping rate,” while to a lesser degree with the “biosample type” and “collection year.” RNA PCs 2-4 correlate with “CM status” (Pearson’s R^2^ > 0.1). The filtered and TMM normalized RNA data shows variation that groups the non-failing and failing (DCM & ICM) samples among the 2^nd^ and 4^th^ PCs (Figure 2A-B). This clustering is likely to explain the combined effects of several genes’ expression profiles and provides evidence for differences between non-failing and failing groups within the TOPCHeF dataset. We show generally across RNA PCs 1-4 that the DCM and ICM samples are overlapping more than with the non-failing group, suggesting an overlap of expression profiles for DCM and ICM (Supplemental Figure 5).

**Figure 2:**
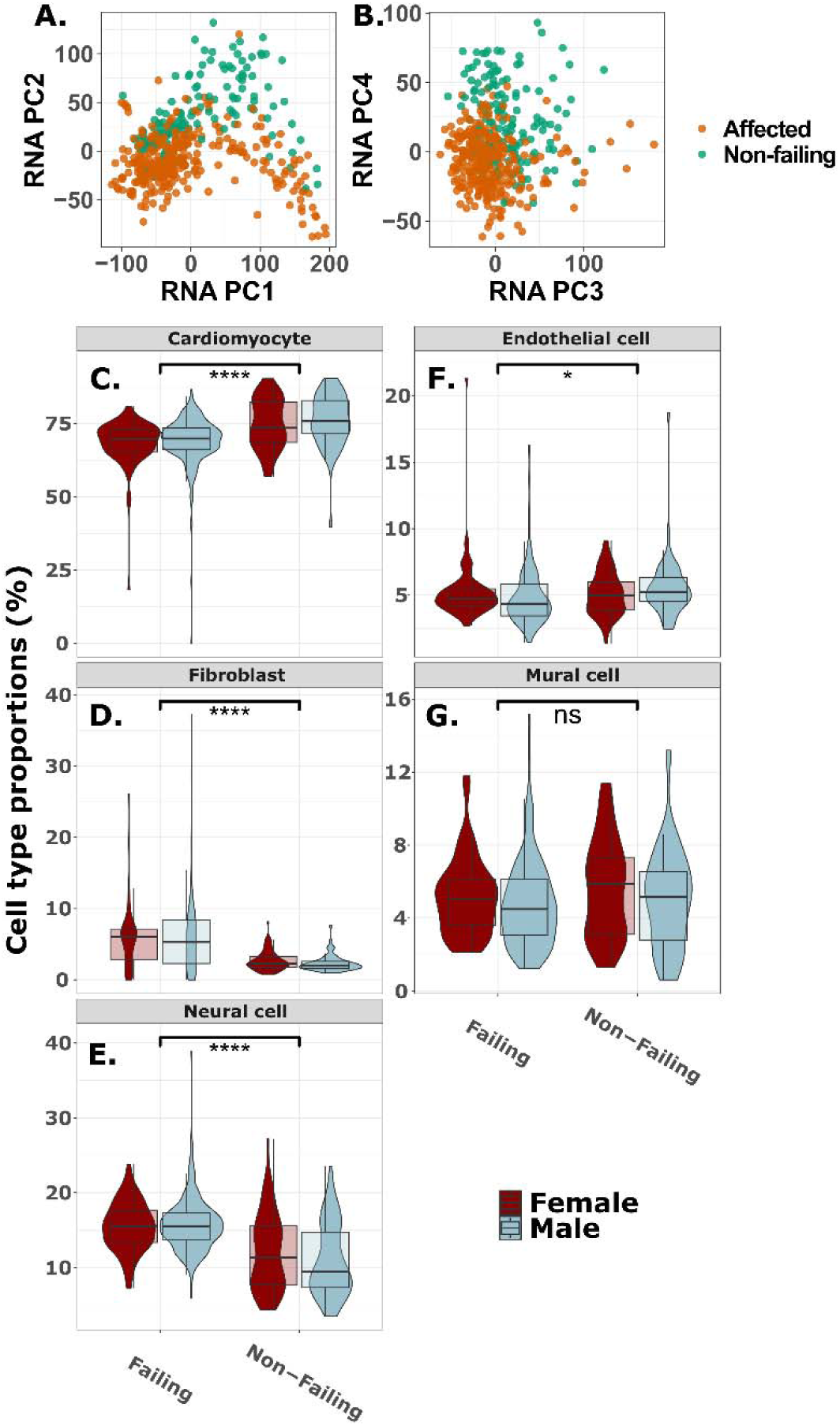
Transcriptional and cell-type variation associated with heart failure. **A.** RNA PCs 1-2 and **B.** RNA PCs 2-3 of the TMM (i.e., trimmed mean of m component) normalized RNA expression for failing and non-failing samples. **C-G.** RNA deconvolution of samples colored by 5 cell-types within left ventricle heart tissue, split by disease group (x-axis) and sex (color) from *MuSiC v2*. Each cell-type is tested with the student’s t-test between group averages of failing and non-failing. **C.** Cardiomyocytes, **D.** endothelial cells, **E.** fibroblasts, **F.** mural cells, and **G.** neural cells. The following symbols represent *p*-values: ns: *p* > 0.05; *: *p* ≤ 0.05; **: *p* ≤ 0.01; ***: *p* ≤ 0.001; ****: *p* ≤ 0.0001.

An additional source of variation in bulk RNAseq experiments can be attributed to differences in cell-type proportions included in the final tissue libraries. We estimated several common cardiac tissue cell-types using the scRNAseq Heart Atlas v2 dataset (Litviňuková et al., 2020) and *MuSiC2*. Estimated deconvolution of the bulk RNA-seq data indicated that most samples were composed primarily of ventricular cardiomyocytes (50–80%), with no significant overall differences between male and female samples (Welch’s two-sample t-test; *t* = 1.5, *df* = 288, *p* = 0.14; Figure 2C). However, when stratified by disease status (DCM & ICM versus non-failing), significant differences emerged. In females, DCM samples showed significantly lower cardiomyocyte proportions compared to the non-failing cohort (*t* = −4.3, *df* = 98, *p* = 4.15 × 10^−5^; Figure 2B), as did ICM samples (*t* = −3.13, *df* = 59, *p* = 0.0027; Figure 2C). These differences were even more pronounced in males, where cardiomyocyte proportions were significantly reduced in both DCM (*t* = −5.74, *df* = 115, *p* = 7.9 × 10^−8^; Figure 2C) and ICM (*t* = −4.78, *df* = 92, *p* = 6.9 × 10^−6^; Figure 2C) samples relative to the non-failing cohort.

Outside of sex-specific differences, the deconvolution results show significant differences in the proportions of cardiomyocytes between the combined HF group (DCM & ICM) and non-failure cohorts (*t* = −7.4, *df* = 232, *p* = 2.8 x 10^−12^; Figure 2C). We also show significant differences in the proportions of fibroblasts (*t* = 12, *df* = 453, *p* < 2.2 × 10^−16^; Figure 2D), neural cells (*t* = 8.9, *df* = 184, *p* = 5.4 × 10^−16^; Figure 2E), and endothelial cells (*t* = −2.17, *df* = 253, *p* = 0.031; Figure 2F) between the affected and non-failing cohorts. There were no significant differences between the diseased cohorts and the non-failing groups within mural cells (*t* = − 1.72, *df* = 218, *p* = 0.087; Figure 2G).

### Heart failure expression-QTL and colocalization with DCM GWAS

We found 10,241 genes with significant cis-eVariants (permutation *p*-value < 0.05) of which, 47% are upstream and downstream region variants, 27.5% are intronic, and 11% are intergenic regions, the remaining ∼15% of variants compose missense (1%), synonymous (1%), and 5’ and 3’ untranslated region (UTR) variants (8%) among others. The mean slope of the eGenes is 0.016 with a 95% quantile of –0.81 and 0.93 and we find that the mean absolute distance away from the TSS is 245,198 bps for cis-eQTL. Around 75% of eGenes (N = 7,691) have a fine-mapped variant in the 95% credible set (when using Susie-TensorQTL), and of these a majority of eGenes have more than one eVariant, while ∼40% have more than 10 fine-mapped variants.

We chose to perform colocalization under a single causal variant assumption (Giambartolomei et al., 2014) to prioritize genes and variants associated with HF, specifically targeting DCM as there are large effect Mendelian alleles that can help explain disease state (McNally & Mestroni, 2017). Jurgens et al., (2024) GWAS summary statistics were used to run single-locus colocalization for each gene (Figure 3A). We found 21 colocalized genes, identified as those with posterior probability H4 (PP.H4) > 0.8 (Figure 3A-B), 16 of which have flipped effect signs between GWAS and eQTL (either positive in GWAS and negative for eQTL and vice versa), 6 with corresponding positive, and 6 corresponding negative effects between GWAS and eQTL (Table 2). The gene *SKI* had the highest PP.H4 of 0.997, with a strong eQTL effect size (β = 0.227 [SE: 0.032], standard deviation units of expression per effect allele), and has been implicated in several cancers due to it being a nuclear proto-oncogene (Luo, 2004), and *SKI* plays a role in the development of cardiac myofibroblasts (Cunnington et al., 2013). The *SKI* sentinel eQTL variant is *rs2503715* (i.e., chr1:2,212,668), a regulatory region variant 15.6 kpbs upstream of *SKI* within *FAAP20* (*FA Core Complex Associated Protein 20*). Here, we define the term “sentinel variant” as the tested mutation with the lowest (i.e., most significant) GWAS *p-*value within each nominal QTL window. The second highest PP.H4 was for *ACTN2* (*Alpha-actinin-2*), a protein expressed in both skeletal and cardiac muscles and functions to anchor myofibrillar actin thin filaments and titin to Z-discs and has been linked with DCM (Mohapatra et al., 2003). The effect size for the *ACTN2* cis-eQTL is β = 0.134 [0.03] and the sentinel variant *rs4659701* (i.e., chr1:236,689,867) is an intronic variant within *ACTN2*.

**Figure 3:**
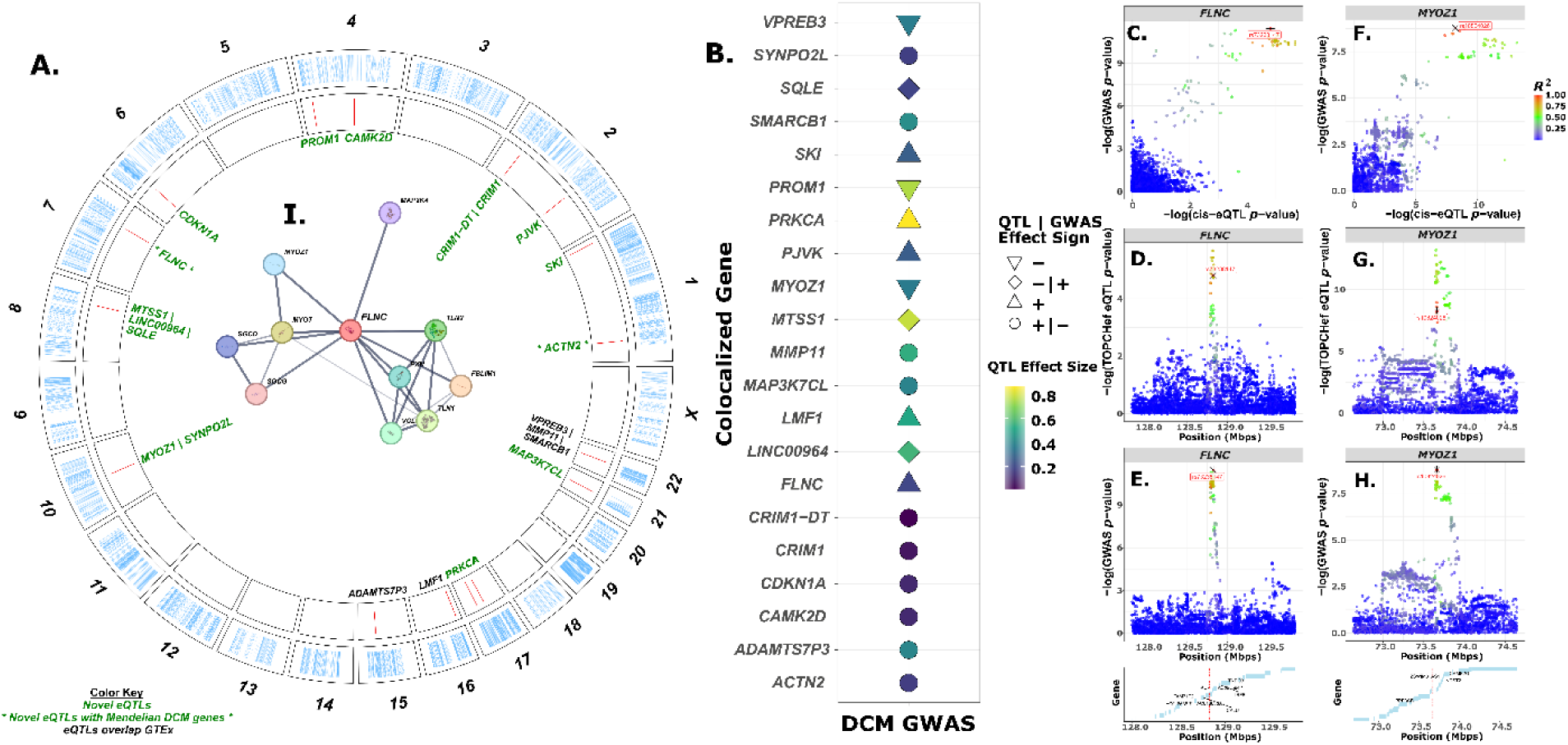
Genome-wide cis-eQTL architecture and colocalization with a large DCM GWAS. **A.** Circos plot showing the genomic distribution of significant cis-eQTL (permutation *p* < 0.05; outer ring) and regions that colocalize with DCM GWAS signals from Jurgens et al. (2024) (PP.H4 > 0.8; inner ring). Genes labeled in green represent loci with no GTEx overlap, and genes in green with an asterisk denote non-overlapping loci that are also known as Mendelian DCM genes. Genes labeled in black indicate an overlap with GTEx. **B.** Colocalized genes identified between cis-eQTL and the DCM GWAS. Point shape indicates whether the eQTL and GWAS effect directions are concordant or discordant, and point color reflects the absolute eQTL effect size. **C–H.** Regional colocalization plots comparing eQTL *p*-values with GWAS association statistics. Panels **C–E** show the *FLNC* locus (±1 Mb from the transcription start site), and panels **F–H** show the *MYOZ1/SYNPO2L* region tested**. I.** Protein–protein interaction (PPI) network from STRING using FLNC as the query gene; edge thickness reflects the confidence of each interaction.

**Table 2:** Colocalized eGenes and their sentinel GWAS SNPs. We identified high confidence colocalized genes (PP.H4 > 0.8) and list sentinel variant annotations and locations within the HG38 assembly. *FLNC* is in bold due to it being a previously identified Mendelian risk gene for DCM diagnosis.

We also tested an *a priori* list of 57 genes known to be involved in various Mendelian DCM cases (Supplemental Table 1). An interesting gene that we found colocalized with the eQTL from this *a priori* list is *FLNC* (*Filamin-C*), a protein coding gene that participates in anchoring of other membrane proteins within the cytoskeleton, is expressed in striated muscle (Thompson et al., 2000), and has been associated with DCM and is used for genetic screening of heart illness in the clinic (Verdonschot et al., 2020). We show that *FLNC* has positive effect sizes in both cis-eQTL and GWAS summary statistics (Figure 3B-E). The lead sentinel variant is *rs73238147* (i.e, chr7:128,829,863), a regulatory region variant 543 bps upstream of *FLNC* (Figure 3C-E). There are 40 fine-mapped variants that are within the 95% credible sets for *FLNC*.

Outside of the known Mendelian genes, the colocalized regions generally show moderate effect sizes on gene expression (β ≥ 0.15) and there are several eVariants that correspond to multiple nearby genes, as is the case for 4 of the tested genes (Figure 3A-B). One such region encompasses both *MYOZ1* and *SYNPO2L* genes that share a highly significant GWAS variant (*p* < 5 × 10^−8^). *MYOZ1* is a *myozenin 1* protein primarily expressed in skeletal muscle and helps properly tether the sarcomere, and has been implicated in HF (Wass et al., 2024). We show that a 3’ UTR variant, *rs10824026* (i.e., chr10:73,646,383), is the eVariant for both *MYZO1* and *SYNPO2L* (Figure 3F-H). In *MYOZ1*, the eQTL effect size is large and concordant in sign with GWAS (β = −0.41 [0.05], *p* = 6.1 × 10^−14^) and has 14 fine-mapped variants. *SYNPO2L* is predicted to be involved with stress fiber assembly and sarcomere organization and has a sentinel eQTL with a β = 0.16 [0.03] and is flipped in sign with GWAS. *MYOZ1* and *SYNPO2L* are distal to one another ∼13 kbps from each other’s TSS and could be implicated in each other’s regulation.

Interestingly, we identified 44 significant trans-eQTL associated with *MYOZ1* (*p* < 5 × 10^−9^). Notably, 19 of these (42.2%) are localized within a region of chromosome 6 (chr6:166,122,098–166,166,769) overlapping the T-box transcription factor T (*TBXT*) gene, and all exhibit positive effect directions. *TBXT* has been implicated with multiple cancers (Palena et al., 2007) and appears to be important with the evolution of tail loss and walking in hominins (Xia et al., 2024). This region appears to represent a trans-eQTL hotspot, as *TBXT* is associated with several hundred significant trans-eQTL involving other genomic regions, consistent with the broad coregulatory influence typical of transcription factors. In contrast, *SYNPO2L* does not have any significant trans-eQTL or evidence of hotspot activity. Beyond these two colocalized examples, *SMARCB1* has five significant trans-eQTL, and *SKI* has one.

Notably, *MYOZ1* is known to interact with *FLNC* and *ACTN2* (Liu et al., 2021). When we estimated protein-protein interactions (PPI) on the amino acid sequence of *MYOZ1*, results showed that *ACTN2* is the most likely binding partner (PPI score = 0.998) and *FLNC* is the 4^th^ with a score of 0.98, both of which are supported by experimental evidence. A reciprocal PPI search with *FLNC* showed that *MYOZ1, MYOZ3, and SMUF2* are all binding partner proteins (score > 0.98; Figure 3I), so the interaction between *FLNC* and *MYOZ1* is nuanced and will be important to further research into the regulatory variation landscape.

### Heart failure splicing-QTL landscape and colocalization with DCM GWAS reveals 17 colocalized splicing regions

Splicing-QTL (sQTL) were tested across 80,750 total isoforms at both *cis* and *trans* scans, consisting of 13,060 genes after quality control and filtering (See Materials and Methods). There were 13,540 significant (permutation *p*-value < 0.05) cis-sVariants across 5,584 genes. We focus further analyses on sQTL if they are both significant DCM GWAS loci (*p* < 5 × 10^−8^) and are colocalized with the Jurgens et al., (2024) DCM GWAS at a PP.H4 ≥ 0.8. We identify four sVariants present across four genes and 17 total isoforms (Figure 4A; Supplemental Table 2). The four colocalized sVariants are sentinel variants for an average of four isoforms each, ranging from influencing one to nine distinct isoforms. We show an overlap with the eQTL colocalization results (see above) in that *SYNPO2L* has three isoforms (splice ID: chr10:73,648,879:73,649,811:clu_9644_-; Figure 4B-C) which are colocalized under a singular sVariant, *rs10824026* (chr10:73,661,450; PP.H4 = 0.97; *p* = 6.14 × 10^−25^; β = −0.62 [0.06]). This intronic variant lies within *SYNPO2L-AS1* and has been implicated with atrial fibrillation (Ellinor et al., 2012). *rs10824026* is the sVariant for three isoforms of *SYNPO2L* with high PP.H4 (> 0.93).

**Figure 4:**
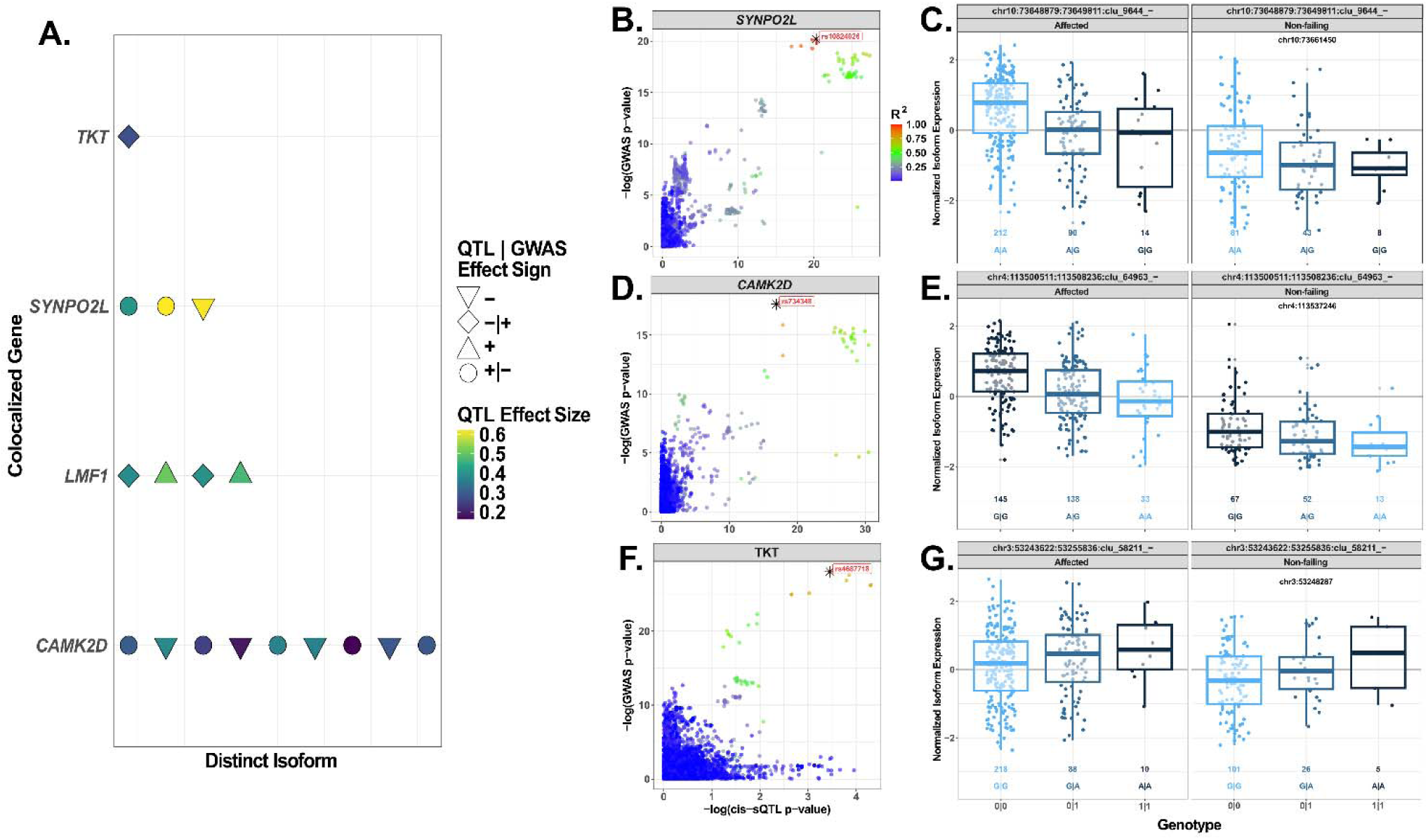
Colocalized cis-sQTL in protein-coding genes. **A.** Summary of colocalized isoforms for four protein-coding genes (*TKT, SYNPO2L, LMF1, CAMK2D*). Each point represents a distinct isoform, with point shape indicating concordance or discordance between sQTL and GWAS effect directions, and color indicating the magnitude of the sQTL effect size. **B, D, F.** Regional colocalization plots for *SYNPO2L* **B),** *CAMK2D* **D),** and *TKT* **F),** showing −log_10_ GWAS *p*-values plotted against −log_10_ cis-sQTL *p*-values. Points are colored by LD (R²) with the sentinel variant; stars denote lead colocalized variants and their rsIDs. **C, E, G.** Genotype-expression plots for representative colocalized isoforms of *SYNPO2L* **C),** *LMF1* **E),** and *TKT* **G).** Boxplots show normalized isoform expression across individuals stratified by genotype: reference homozygotes (0|0), heterozygotes (1|0), and individuals carrying two effect alleles (1|1), displayed separately for affected and non-failing hearts.

The most abundant colocalized cis-sVariant was *rs734348* (Figure 4D-E; chr4:113,537,246; PP.H4 = 0.96, *p* = 1.09 × 10^−10^; β = −0.19 [0.04]), an intronic variant within *CAMK2D* (spliceID: chr4:113,500,511:113,508,236:clu_64963_-:) that is associated with nine different isoforms of the same gene. Again, *rs734348* is of special interest in that it is also the colocalized sentinel eVariant within *CAMK2D* (Figure 3A-B) and is therefore associated with expression and splicing variation. Another notable colocalized isoform is with *TKT* (Figure 4F-G), where the sVariant is *rs62256002* (chr3:53,248,974; PP.H4 = 0.88), an intronic variant affiliated with electrocardiogram morphology amplitudes (Radhakrishnan et al., 2023; Verweij et al., 2020). The sentinel variant with *TKT* (chr3:53,243,622:53,255,836:clu_58211_-) is the sVariant, *rs4687718* (chr3:53,248,287), an intronic variant that has 13 phenotypes associated with it on the Ensembl browser, among which the QRS duration is supported (Prins et al., 2018). Finally, we show that the following sVariant, *rs9889137*, (chr16:918717; PP.H4 = 0.91; *p* = 4.3 x 10^−10^, β = −0.39 [0.07]) is colocalized with three isoforms of the *LMF1* gene (lipase maturation factor-1), a protein-coding gene that plays a critical role in the maturation of lipases (Figure 4A). *rs9889137* has been reported as an associated mutation in type two diabetes progression in a recent GWAS (Suzuki et al., 2024). But *rs9889137* is of specific interest because it is also the colocalized eVariant for *LMF1* (Figure 3A-B), so it shares association with both splicing and expression changes.

The trans-sQTL analysis revealed 14,296 significant trans-sVariants across the genome (α = 5 × 10⁻L/80,750), corresponding to 11,216 unique trans-sVariants that influence 769 total genes. Several genes, including *C14orf180* and *HSD17B7* each harbor more than 1,000 trans-sVariants, highlighting their extensive regulatory connections. We did not identify any colocalized cis-sQTL that also overlap with trans-sQTL; however, we did find overlaps with cis-eQTL. One notable trans-sVariant, *rs146454235* (chr15:69179201), is an intronic variant within *GLCE* associated with *TMEM182* (chr2:102,762,349:102,762,587:clu_49864_+; *p* = 9.71 x 10^−16^; β = −1.11 [0.13]). Additionally, *VPS11* has 3 intergenic sVariants (chr6:14434109, chr6:14,435,028, chr6:14,436,917) that influence the same isoform (chr11:119,073,951:119,076,897:clu_16847_+), all showing negative β between –1.1 and –1.33. Finally, *NMB* has an sVariant, *rs16958020* (chr16:82,654,393; chr15:84,657,348:84,657,996:clu_28884_-; *p* = 3.77 x 10^−13^; β = −1.43 [0.19]), which is an intronic variant within *CDH13*.

### Replication and Functional Relevance of TOPCHeF QTL

To evaluate the reproducibility and biological relevance of our findings, we compared TOPCHeF QTL against external reference datasets and disease-associated loci. Replication in independent cohorts provides a critical benchmark for the robustness of QTL discovery, while overlapping with GWAS loci highlights potential mechanisms by which regulatory variation contributes to disease.

We first assessed replication with the GTEx LV dataset, one of the largest resources for transcriptomic regulation in non-diseased human hearts. Here, 36% of significant TOPCHeF eQTL overlapped with GTEx LV cis-eQTL, increasing to 51% when considering 95% credible sets (Supplementary Figure 6). For splicing, 24.7% of TOPCHeF cis-sQTL overlapped with GTEx LV sQTL, rising to 46.5% with credible sets (Supplementary Figure 6). We also evaluated the Myocardial Applied Genomics Network (MAGNet) resource, which includes both HF and control non-failing samples. In failing hearts, 24.5% of eQTL and 23.3% of sQTL overlapped with TOPCHeF signals, while in non-failing controls, the overlap was similar at 24.5% for eQTL and 26.1% for sQTL (Supplementary Figure 6).

Next, we asked whether TOPCHeF QTL simply overlap with loci associated with DCM. Among the reported 780 genome-wide significant (*p* < 5 × 10^⁻^□) DCM GWAS sites, we observed that 13.2% (N = 103) of eQTL 95% credible sets overlapped with disease loci. sQTL showed a slightly higher overlap of 14.7% (N = 115) across the 2,585,070 nominally significant sites (Supplementary Figure 6). Together, these analyses demonstrate that TOPCHeF QTL not only replicate in independent heart transcriptomic datasets but also intersect with genetic risk loci for DCM, underscoring their potential role in mediating disease-relevant regulatory mechanisms.

## Discussion

This study presents the Trans-Omics for Precision Medicine in Congestive Heart Failure (TOPCHeF) dataset as a comprehensive quantitative trait loci (QTL) resource that integrates whole-genome and transcriptome sequencing from failing (DCM & ICM) and control non-failing human hearts. Through eQTL and sQTL mapping, coupled with colocalization of DCM GWAS loci, we uncovered thousands of cis-regulatory effects with over 10,000 significant eQTL and ∼8,600 sQTL, with over 30 genes colocalizing with known DCM risk loci. Our findings demonstrate that integrating genomic and transcriptomic data derived directly from the failing human heart LV that can identify both known and novel molecular mechanisms of HF. By connecting common genetic variation to transcriptional and splicing regulation, TOPCHeF serves as a reference for both precision medicine and exploratory genetics research approaches in cardiomyopathy research.

### Expression and splicing regulatory variation associated with dilated cardiomyopathy

Our results establish that several well-characterized Mendelian DCM genes: like *FLNC* and *ACTN2* harbor cis-regulatory variants, called eVariants, that colocalize with DCM GWAS loci. These results showcase the robustness of our QTL mapping and confirm the sensitivity of the TOPCHeF dataset to detect known regulatory signals within structural genes implicated in sarcomere organization and cytoskeletal integrity. Importantly, we also identified non-Mendelian loci such as *MYOZ1*, *SYNPO2L*, and *SKI*, which influence expression and for the case of *SYNPO2L*, both expression and splicing regulation in the failing heart. These genes may represent mechanistic intermediates connecting common genetic risk to molecular dysfunction in DCM. For instance, *SKI* emerged as the top colocalized eGene (Figure 3B; PP.H4 = 0.997): a proto-oncogene known to modulate the cardiac fibroblast phenotype (Cunnington et al., 2013). This finding implicates SKI-mediated fibrotic pathways in DCM. Similarly, we confirmed colocalization at Mendelian DCM loci such as *ACTN2* and *FLNC*, reflecting the well-known role of Z-disc/sarcomeric proteins in cardiomyocyte structure (Jurgens et al., 2024). And the result that *MYOZ1* and *SYNPO2L* share regulatory variants within a close genomic interval (∼3 kbps from gene start to stop) suggests a connected transcriptional architecture linking sarcomeric scaffolding and signaling pathways in the failing heart (Wass et al., 2024).

Beyond validating Mendelian DCM genes, our integration of eQTL and sQTL underscores the value of exploring both expression and splicing mechanisms in parallel. Many studies have begun to focus on QTL discoveries and provide insights into how genetic variants may modulate transcript variety within the left-ventricle. Our splicing QTL analysis further broadened the molecular landscape of DCM. We identified cis-sQTL variants that affect the processing of genes involved in calcium signaling and metabolism. Notably, an sVariant in *CAMK2D* colocalizes with DCM (PP.H4 = 0.96), influences multiple *CAMK2D* isoforms, and is associated with expression changes (Figure 3B; Figure 4A, D-E). *CAMK2D* encodes *CaMKII*δ, a key kinase in cardiac calcium handling, and its dysregulation is known to affect heart function (Braz et al., 2004; Jurgens et al., 2024; Sutanto et al., 2020). Alternative splicing of *CAMK2D* could modulate an aspect of calcium-dependent signaling in failing hearts. We also found sVariants affecting isoforms of *TKT* and *LMF1*, enzymes involved in metabolic pathways. *TKT* (transketolase) participates in the pentose phosphate pathway and has been linked to cardiac electrophysiological traits (Wang et al., 2024). Additionally, *TKT* was found to play an intrinsic role in the ischemic HF of mice models (Péterfy et al., 2007). *LMF1* (lipase maturation factor) is a chaperone for lipase enzymes and has been implicated with large-effect mutations increasing the risk of hypertriglyceridemia (Bedoya et al., 2025; Péterfy et al., 2007). *LMF1*, like *SYNPO2L* and *CAMK2D*, share the same e– and sVariant, making it special in that the sentinel mutation is influencing both splicing and expression of *LMF1* in cis (upstream ∼32 kbps from TSS). The finding that there are sQTL within genes involved in cytoskeletal and calcium signaling processes may suggest that disease-associated variation may often act through splicing perturbations. This has important implications for functional validation studies, as disease mechanisms may depend on isoform-specific alterations that are not captured by conventional gene-level analyses.

Overlap replication with GTEx and MAGNet datasets confirm that a substantial fraction of cis-regulatory signals is reproducible across independent cohorts (25 – 50%; Supplemental Figure 6), while the presence of unique QTL in TOPCHeF underscores the added value of sampling diseased tissues. Many of the QTL that are not observed in GTEx and MAGNet could suggest that the regulatory architecture of the failing heart diverges from that of non-diseased LV, as revealed by the RNA PCA (Figure 2A-B) and covariate correlation results (Supplemental Figure 5). This observation aligns with prior evidence that disease-specific environments, such as inflammatory signaling, altered chromatin states (Tan et al., 2020), and metabolic stress can unmask regulatory variation otherwise hidden under homeostatic conditions (Revathi Venkateswaran et al., 2024). Thus, TOPCHeF fills a critical gap between large-scale population transcriptomic resources and smaller disease-focused studies by offering a well-powered and tissue-relevant dataset for exploring regulatory variation in the failing heart.

### Limitations and future work

While the TOPCHeF dataset is among the largest for left-ventricles, sample size still limits power for detecting rare variants (MAF ≤ 0.05) and trans-QTL, which typically exhibit smaller effect sizes on a similar scale to GTEx (The GTEx Consortium et al., 2020). Rare variants (MAF < 0.01) are likely to be underpowered in this dataset. For context, the failing heart group includes 316 individuals, and the non-failing group includes 132, corresponding to a minimum observable allele frequency of 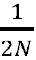 = 0.0016 and 0.0038, respectively. The tissue samples from DCM and ICM were collected from advanced, end-stage HF and from an ancestrally homogenous population making inferences on early or moderate stages of HF and on the effects of non-European ancestry elusive.

While deconvolution methods help mitigate confounding by cell-type composition, bulk RNA-seq datasets can be somewhat heterogeneous across cellular compositions. Our comparative analysis of cell-type proportions using *MuSiC2* did not reveal large variance within groups and within sex, still, there are fluctuations in the range of cellular proportions (Figure 2C-E). Future work integrating single-nucleus transcriptomics and spatial omics from the same individuals could disentangle cell-specific regulatory mechanisms and cell-type interacting QTL (e.g., iQTL; Kasela et al., 2024). This is especially interesting because it could help uncover possible regulatory variation with cardiomyocytes between the failing and non-failing samples (Figure 2C-E).

Lastly, while colocalization provides statistical evidence of shared causal variants, experimental perturbation and CRISPR-based validation will be necessary to confirm direct mechanistic links between genotype and phenotype within human heart tissue or cardiac induced pluripotent stem cells (i.e., hiPSC-CMs; Deicher & Seeger, 2021). Collectively, the major value of TOPCHeF is that it will aid in the discovery of genetic effects that generalize across multiple forms of HF (i.e., DMC, ICM, and other cardiomyopathies). These attributes make the dataset an ideal reference for integrating additional molecular modalities (as seen in TOPMed, Orchard et al., 2025), such as chromatin accessibility (ATAC-seq), methylation, proteomics (e.g., pQTL), and metabolomics to develop a more complete system-level understanding of HF (Rasooly et al., 2025) from the evolving sample biobank.

## Conclusion

The TOPCHeF project establishes a foundational resource for understanding the genetic regulation of the failing human heart. By leveraging genomic, transcriptomic, and splicing analyses in a large, clinically characterized cohort, we bridge the gap between population-scale reference datasets and disease-focused molecular investigations. The discovery of colocalized regulatory variants in both known (*FLNC, ACTN2*) and novel cardiomyopathy genes illustrates how integrating multi-omic data from diseased tissue can refine our interpretation of GWAS findings and identify actionable targets for precision medicine. Beyond its immediate findings, the availability of the TOPCHeF dataset will enable continued discovery across diverse cardiac phenotypes.

### Code Availability

Source code for the analyses, pipeline, and figures are available here: https://github.com/connor122721/nf-eqtls.

### Data Availability

Individual level controlled-access data for the TOPCHeF DNA and RNA sequencing are available through dbGAP (dbGaP ID:phs002038). TOPCHeF QTL summary statistics are available for download on Zenodo (10.5281/zenodo.17932667).

## Acknowledgements and Funding Information

This research was supported by R01-HL170012 (AM and MRGT). Molecular data for the Trans-Omics for Precision Medicine (TOPMed) program was supported by the National Heart, Lung and Blood Institute (NHLBI) award (X01 HL139403). Whole genome sequencing and RNASeq for “NHLBI TOPMed: Trans-Omics Analysis for Congestive Heart Failure (TOPCHeF)” (phs002038.v1.p1) was performed at the Broad Institute’s Genomics Platform (HHSN268201600034I). Core support including centralized genomic read mapping and genotype calling, along with variant quality metrics and filtering were provided by the TOPMed Informatics Research Center (3R01HL-117626-02S1; contract HHSN268201800002I). Core support including phenotype harmonization, data management, sample-identity QC, and general program coordination were provided by the TOPMed Data Coordinating Center (R01HL-120393; U01HL-120393; contract HHSN268201800001I). Study support for the Colorado Cardiac Tissue Bank comes, in part, from the University of Colorado Division of Cardiology. REDCap was provided by National Institutes of Health/NCATS Colorado CTSA Grant Number UM1 TR004399. Infrastructure for the CHARGE Consortium is supported in part by the National Heart, Lung, and Blood Institute (NHLBI) grant R01HL105756.

We sincerely thank all the patients who donated their heart tissue and data for this work and their commitment to furthering heart failure research for many. We gratefully acknowledge the contributions of Dr. Michael Bristow, MD, PhD, whose insights greatly enriched this work prior to his passing in November 2025. The authors wish to acknowledge members of the Manichaikul lab, including: Wiktor Nisterenko, Xiaowei Hu, Catherine Debban, Brody Receveur, Paula Czarnota, and Sam Park for their discussion and feedback related to this work’s development. We also thank Peter Orchard for advice on QTL mapping. The authors acknowledge Research Computing at the University of Virginia for providing computational resources and technical support that have contributed to the results reported within this publication. URL: https://rc.virginia.edu. Contents are the authors’ sole responsibility and do not necessarily represent official NIH views.

## Supplemental Figures

**Supplemental Figure 1:**
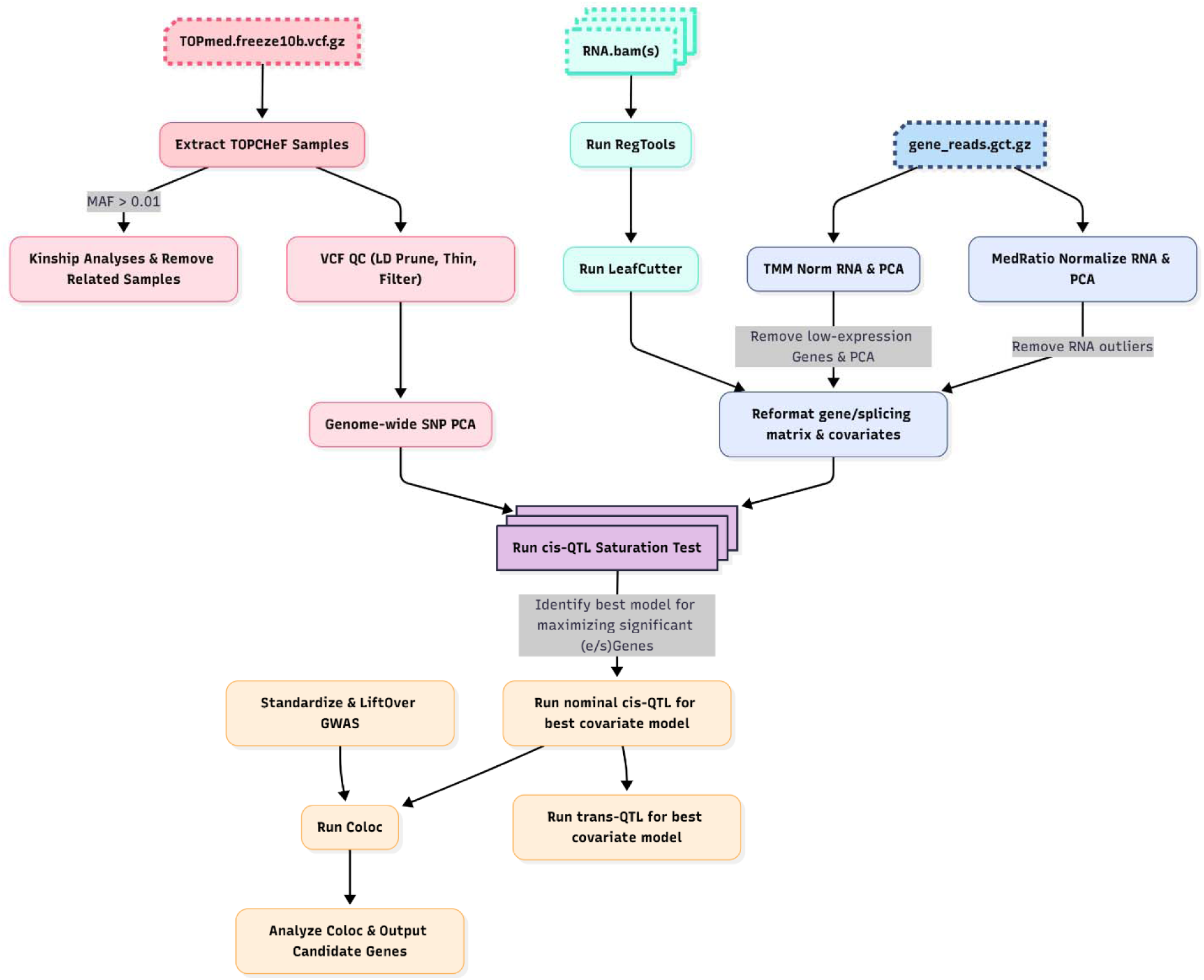
Computational pipeline for the genetic, transcriptomic, and splicing processing and QTL mapping. This pipeline is split between the genetic processing of whole-genome sequencing (pink), RNA-sequencing (blue), and splicing (cyan).

**Supplemental Figure 2:**
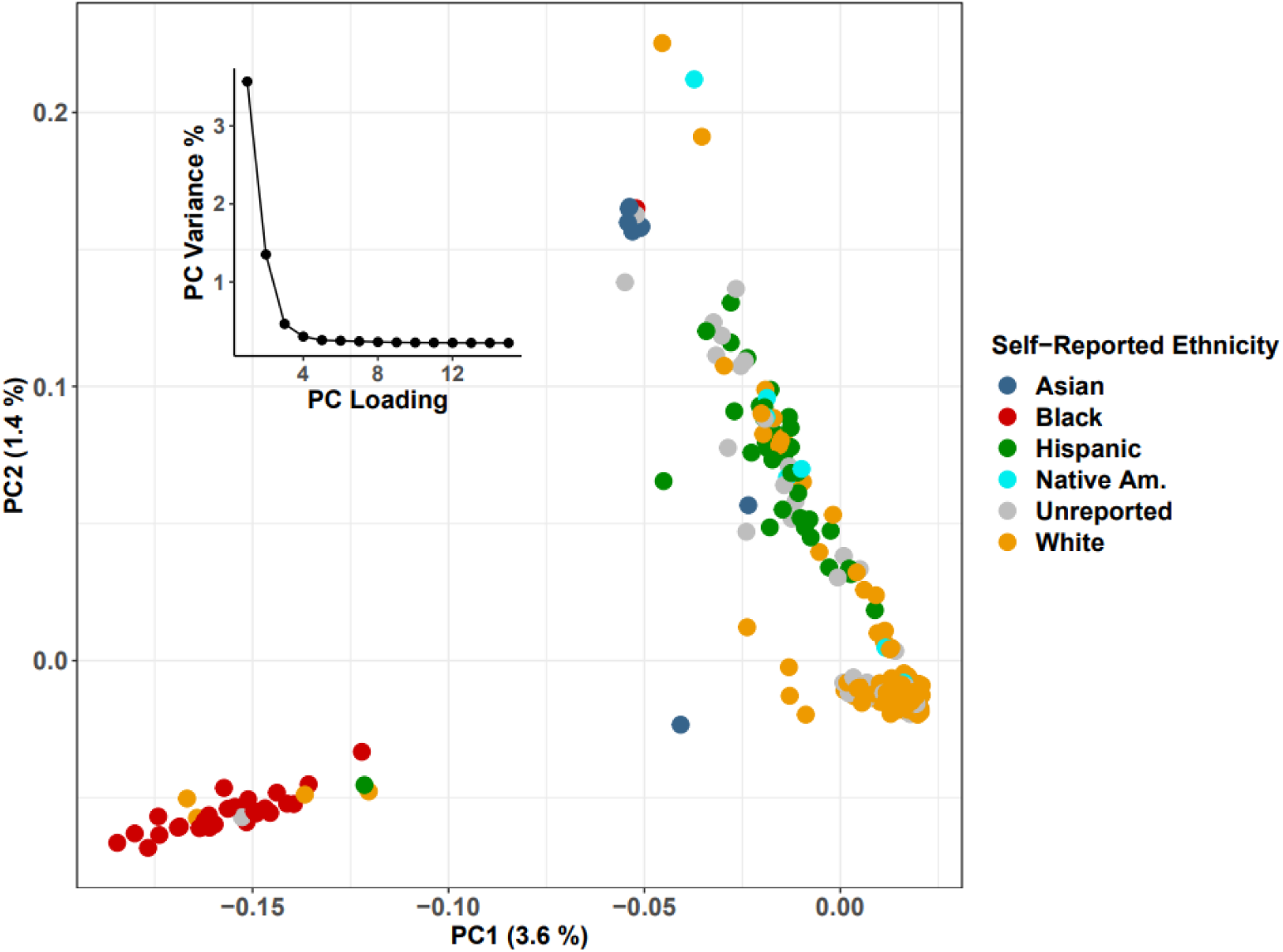
Genetic variation within TOPCHeF. PCA of minor allele frequency filtered (MAF > 0.05) and thinned dataset colored by self-reported ancestry. Inset plot is the proportion of variance explained by each PC loading for the first 15 PCs.

**Supplemental Figure 3:**
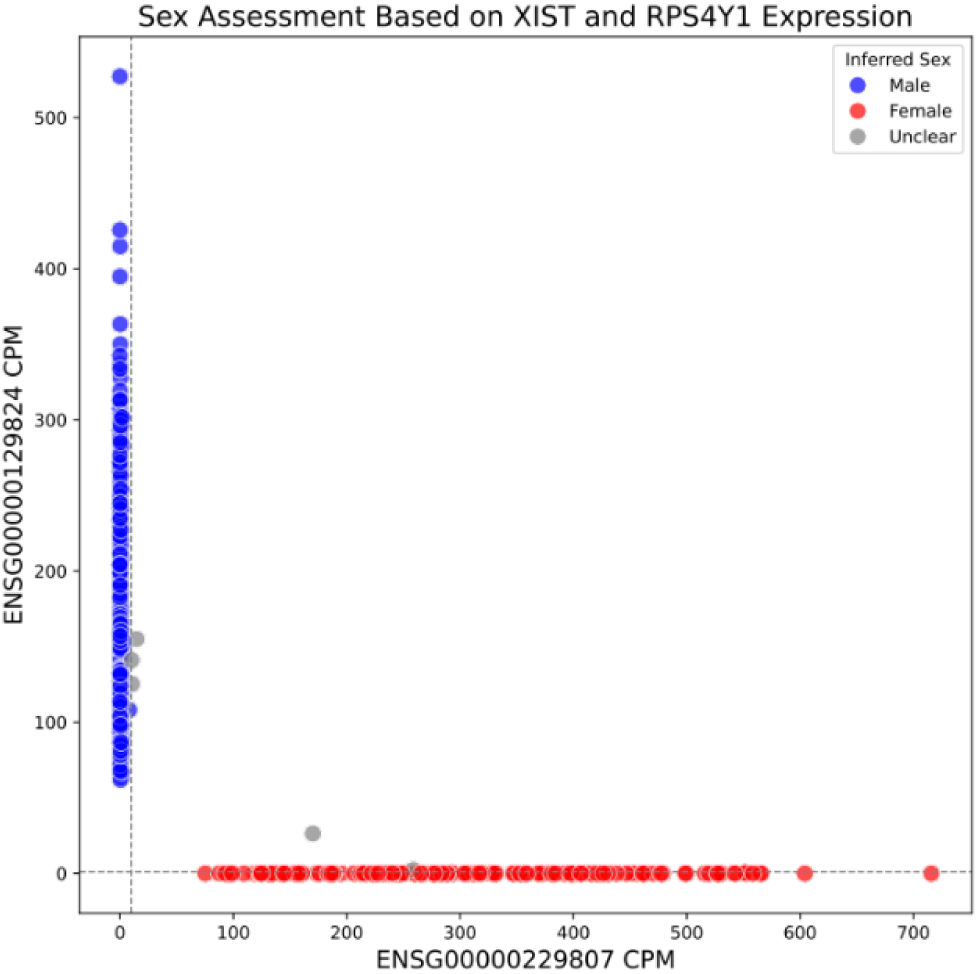
Assigned sex metadata overlaps with sex determination based on XIST and RPS4Y1 expression. Each point is a sample, post-TMM normalization. The three unclear samples were retained because they did not represent clear cases of contamination. ENSG00000229807 (x-axis) is *XIST* and ENSG00000129824 (y-axis) is *RPS4Y1*.

**Supplemental Figure 4:**
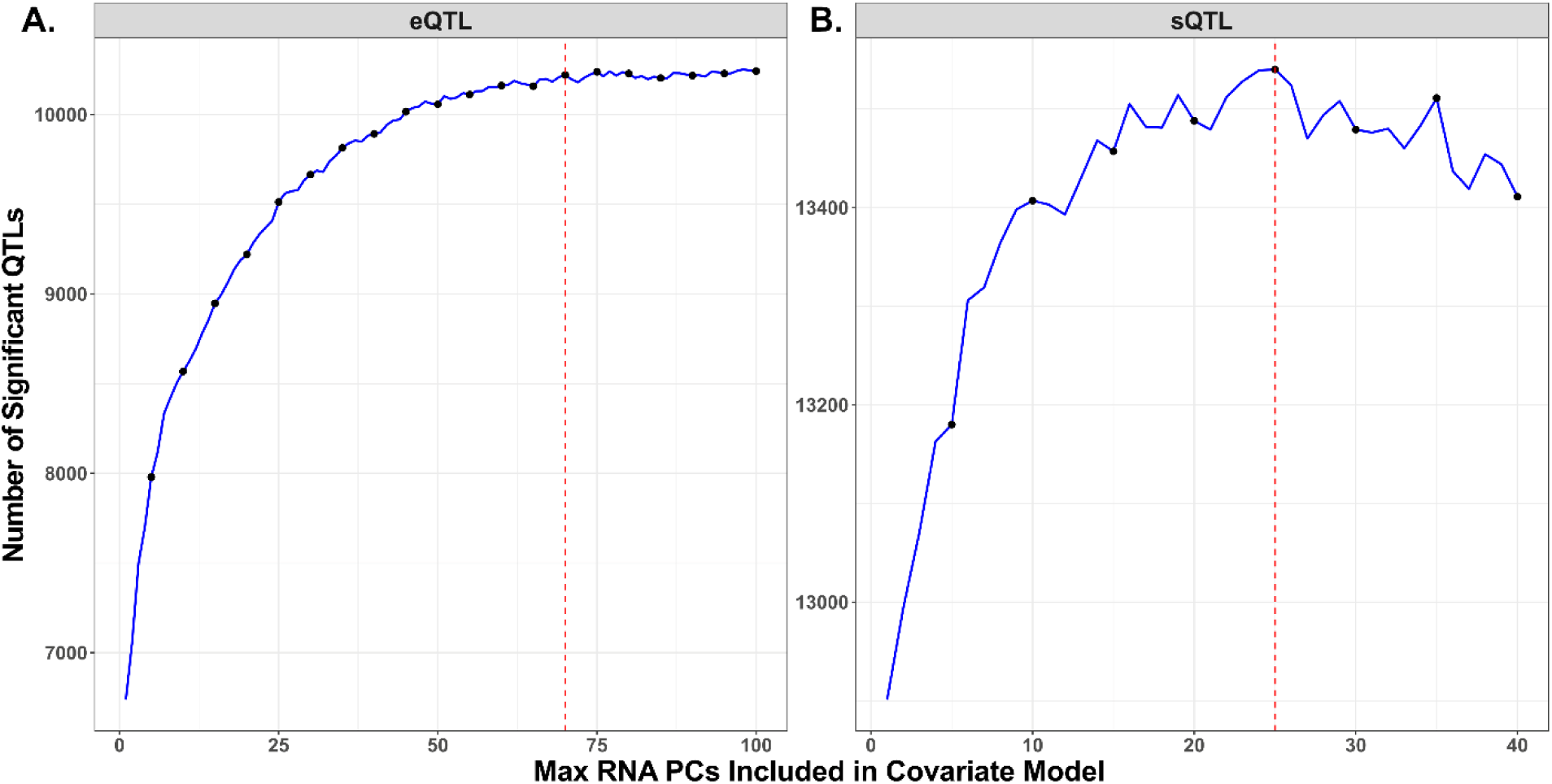
Saturation of cis-QTL are maximized when including several RNA PCs in the covariate models. **A)** Significant eQTL are maximized (i.e., reached a statistical plateau) at RNA PC 70 (red dotted vertical line), we varied the number of maximum PCs included in the covariate model for cis-eQTL. **B)** Significant sQTL are maximized at the RNA PC 25 (red dotted vertical line).

**Supplemental Figure 5:**
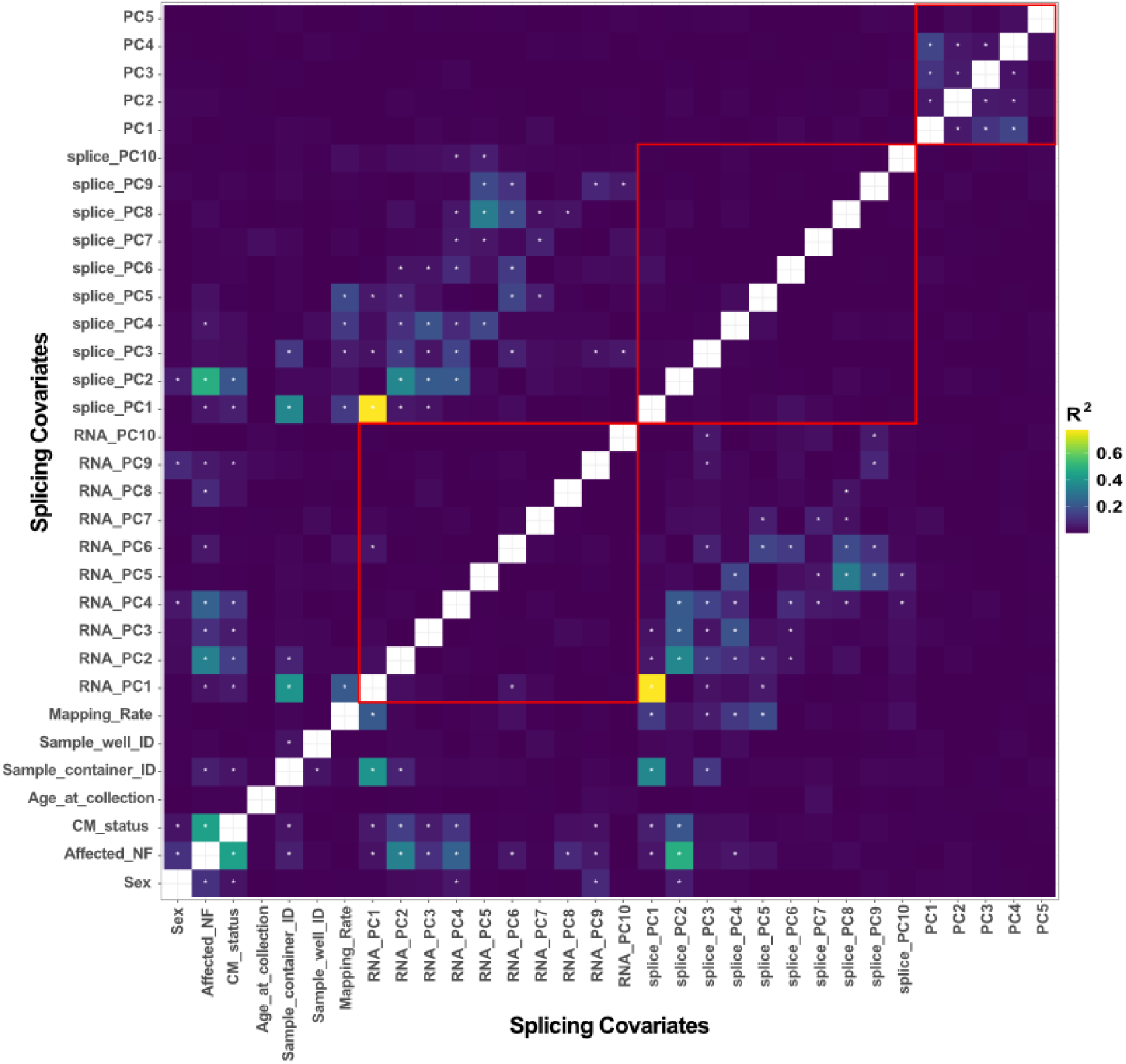
Pairwise correlation of covariates included in eQTL and sQTL mapping. We used the values for every individual and ran Spearman’s correlation statistics to estimate a rank-based measure of association. White stars indicate a significant correlation after multiple testing correction (FDR *p* < 0.05). The red box within the matrix indicates any pairwise expression RNA PCs, splicing RNA PCs, or PCs of ancestry comparisons. We added three covariates (“*Sample_container_ID*”, “*Sample_well_ID*”, and “*Mapping_Rate*”) that are absent from the covariate model for QTL mapping but likely reflect experimental batch effects introduced during sample preparation or RNA extraction.

**Supplemental Figure 6:**
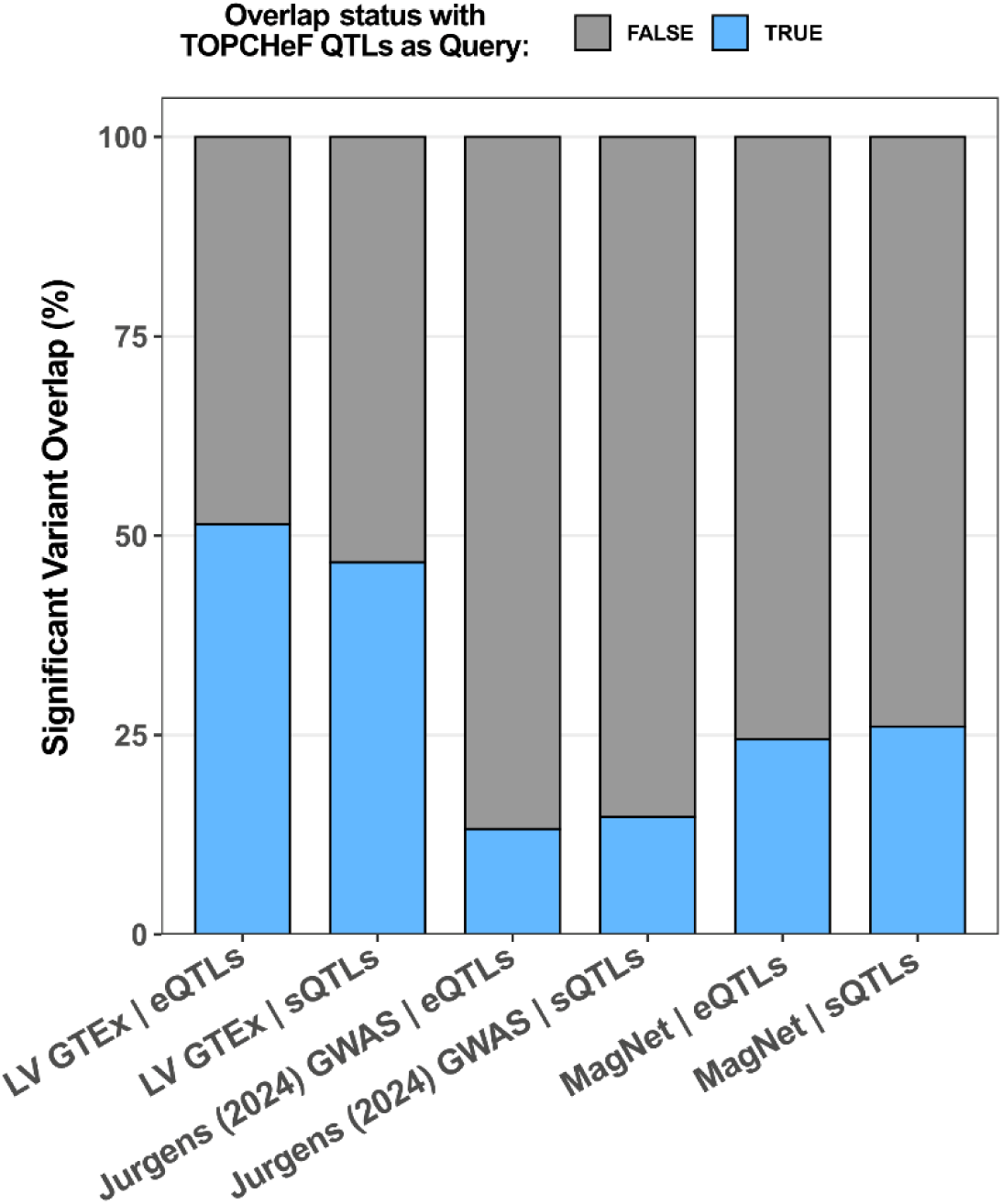
Overlap replication of TOPCHeF QTL with GTEx, GWAS, and MagNet. Proportions of significant eQTL and sQTL that overlap across the three datasets with the significant TOPCHeF QTL.

**Supplemental Table 1:** Candidate Mendelian DCM genes. We identified high confidence colocalized genes (PP.H4 > 0.8) and list sentinel variant annotations and locations within the HG38 assembly.

**Supplemental Table 2:** Colocalized sGenes and their sentinel GWAS SNPs. We identified high confidence colocalized genes (PP.H4 > 0.8) and list sentinel variant annotations and genomic locations.

